# Topology-Aware Optimisation of Vaccination Strategy for Minimising Virus Spreading

**DOI:** 10.1101/2021.10.19.21265198

**Authors:** Pietro Hiram Guzzi, Francesco Petrizzelli, Tommaso Mazza

## Abstract

Vaccination is currently the primary way for mitigating the COVID-19 outbreak without severe lockdown. Despite its importance, the available number of vaccines worldwide is insufficient, and the production rates are hard to be increased in a short time. Therefore, vaccination needs to follow strict prioritization criteria. In this regard, almost all countries have prioritized similar classes of exposed workers: healthcare professionals and the elderly obtaining to maximize the survival of patients and years of life saved. Nevertheless, the virus is currently spreading at high rates, and any prioritization criterion so far adopted did not show to account for the topology of the contact networks. We consider that a network in which nodes are people while the edges represent their contacts may model the virus’s spreading efficiently. In such a model, it is already known that spreading may be efficiently stopped by disconnecting the network, i.e., by vaccinating more *central* or relevant nodes, therefore, eliminating “bridge edges”. Consequently, we introduce such a model and discuss the use of a topology-aware versus an age-based vaccination strategy.

## 1 Scientific Background

The World Health Organization declared on March 2020 the spread of the SARS-CoV-2 virus to be a pandemic [10]. After initial containment measures and the trial of different therapies, it was clear that the primary way to mitigate the spread was to develop a vaccine [11, 15, 14, 17]. Consequently, many pharmaceutical industries developed with an unprecedented speed different vaccines. Currently, four COVID-19 vaccines are approved for use by the FDA and EMA, while other eight are in early testing phases. However, considering the spread of COVID-19 and the production rates, there is a need for developing ad hoc prioritization strategies [4].

The design of a vaccine prioritization strategy is then a crucial challenge in fighting COVID-19 since there is evidence that the production rate will remain insufficient for several months and that COVID-19 has different impacts and transmission rates in other social groups [7, 12]. In particular, COVID-19 showed a higher fatality rate in older people and men [6]. Consequently, different strategies have been discussed and evaluated by the scientific communities to support the governments [9]. These strategies consider as variables the characteristics of vaccines (e.g., the possible variation of the mitigating impact caused by aging), the effect of COVID-19 on social groups (e.g., fatality rates by age, transmission rate by worker class).

Thus, in many countries, these criteria were guided by demographic and social considerations, thereby pushing to prioritize the elderly and healthcare professionals. As demonstrated by Goldstein et al., [9], the vaccination of the oldest enabled to save most lives and years of life. Common sense suggests that a good prioritization scheme should choose the best trade-off between saving the maximum number of lives and the most future life. The mathematical model developed by the authors demonstrated that giving priority to older adults may maximize both effects. Thus the strategy is feasible. In effect, older people’s prioritization was chosen as the main criterium in many countries such as Italy and the US. These countries also gave priority to healthcare professionals, teachers, and caregivers.

Despite the effectiveness of this approach, it has been clear that a vaccine allocation strategy requires the incorporation of a model of transmission and the epidemiological characteristics of the disease among social groups [5, 8, 16]. Buckner et al. developed a framework for the optimal dynamic prioritization of vaccines integrating the transmission dynamics and COVID-19 characteristics into their model. The authors used a compartmental model for modeling COVID-19 spreading. They studied the impact of optimizing three objectives, i.e., minimizing the new cases, the years of life lost (YLL), or the number of deaths. The first contribution of this work is to discuss the fact that no one strategy fits all benefits, i.e., the current limited knowledge of many variables makes it impossible to develop a system that can simultaneously achieve all three objects. Second, the authors demonstrated through three different simulations that the precedent objectives require clearly different strategies. Minimizing mortality (YLL or deaths) requires the vaccination of older essential workers (especially 60 + years). Alternatively, the minimization of the number of cases is achieved by giving priority to essential workers. Authors clearly stated that policies that differentiate and target essential workers in addition to age substantially outperform those considering the age alone.

In [13], Jentsch et al. dealt with the problem of optimizing the vaccination strategy. They demonstrated that a strategy based only on age cannot be optimal compared to a contact-based strategy. They developed a mathematical model fitting on Ontario data and simulated the impact of different vaccination strategies. Their contact strategy was based on the allocation of vaccines according to the relative roles played by different age groups in transmission. This strategy tended to prioritise ages ranging from 15 to 19 years first, 20–59 years second, and gave the least priority to older or younger ages.

The analysis of these approaches stimulates, to the best of our knowledge, two primary considerations: (i) the optimization strategy is dependent on the desired goals, (ii) the integration of the characteristics of the modeling improves the performances, (iii) a dynamic strategy may outperform a static one. Despite this, we reckon that modeling the virus spreading using a classical SEIR model may not be the best choice since some parameters are considered at a global scale, while the spreading involves individuals that are topologically different in terms of their social contacts. In parallel, some previous works such as [1, 19] have demonstrated that the use of a model coming from graph theory may be helpful to describe the spreading. In this way, comprehensive graphs may be derived using nodes, people, and edges of their contacts.

Given such a model, we deem that Graph Theory can suggest a simple optimization scheme to minimize the virus spreading. The spreading of information (or more generally the spreading itself), best conditions for spreading, and the emergence of super-spreaders have been largely investigated in network science [18, 2, 3]. These works discussed the challenge of detecting and suppressing the spread of dangerous viruses, pathogens (misinformation, gossips, or information in general). In [20] authors investigated the impact on the community structure of the network on percolation, simulating the spreading of an epidemic modeled through a SIR model. Authors concluded that spreading within communities is critically related to denseness, while the intercommunity edges are the most important factor in spreading an epidemic, regardless of community size and shape.

In particular, in [21] a study of epidemic spreading using the adjacency matrix of a graph has been proposed, as described in Figure 1. Given a contact matrix without any constraint on its structure and an epidemic modeled using a SIR model defined by the rate of new infections *β* and the rate of recovered people *δ*, it has been demonstrated that there is an upper bound to the epidemic given by:

**Figure 1:**
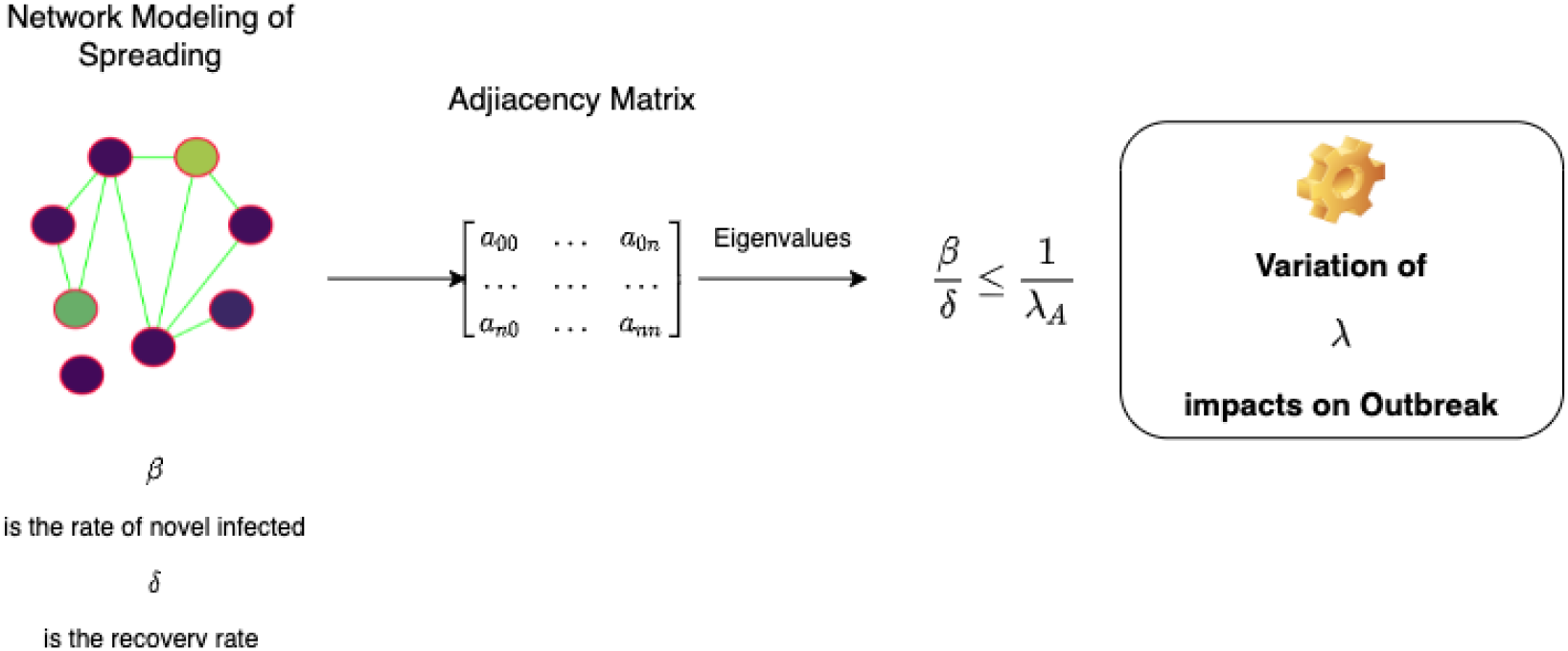
The rationale of the paper. The spreading of the SARS-CoV-2 virus may be modeled using a network whose nodes are people while edges are contacts among them. The spreading may be summarised in a SIR model using two main parameters *β*, and *δ*, representing the rate of novel infected people and the recovery rate. The network may be represented using the adjacency matrix of the resulting graph. It is known that spreading may be contained when following condition holds: 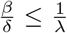, where *λ* is the largest eigenvalue of the adjacency matrix. We hypothesize that the vaccination of an individual is equivalent to the deleting of a node (or equivalently to the deleting of the edge of the contacts).

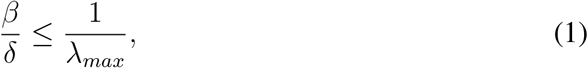

, where *λ*_*max*_ is the largest eigenvalue of the adjacency matrix. Consequently, given the property that

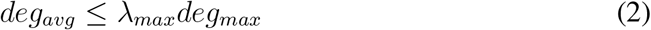

where *deg*_*avg*_ and *deg*_*max*_ are, respectively, the average and the maximum node degree.

Consequently, we propose removing, i.e., to vaccine, the node, i.e., the individuals, with the highest node degree from the graph. In such a way, the value of the largest eigenvalue decreases, and consequently, the left member of the equation increases. We also empirically demonstrate that for a large class of graphs used for modeling social contact, the above-described property also holds for different node centrality measures, i.e., the removal of most central nodes causes the decrease of the value of the largest eigenvalue. The workflow of our empirical demonstration is depicted in Figure 2.

**Figure 2:**
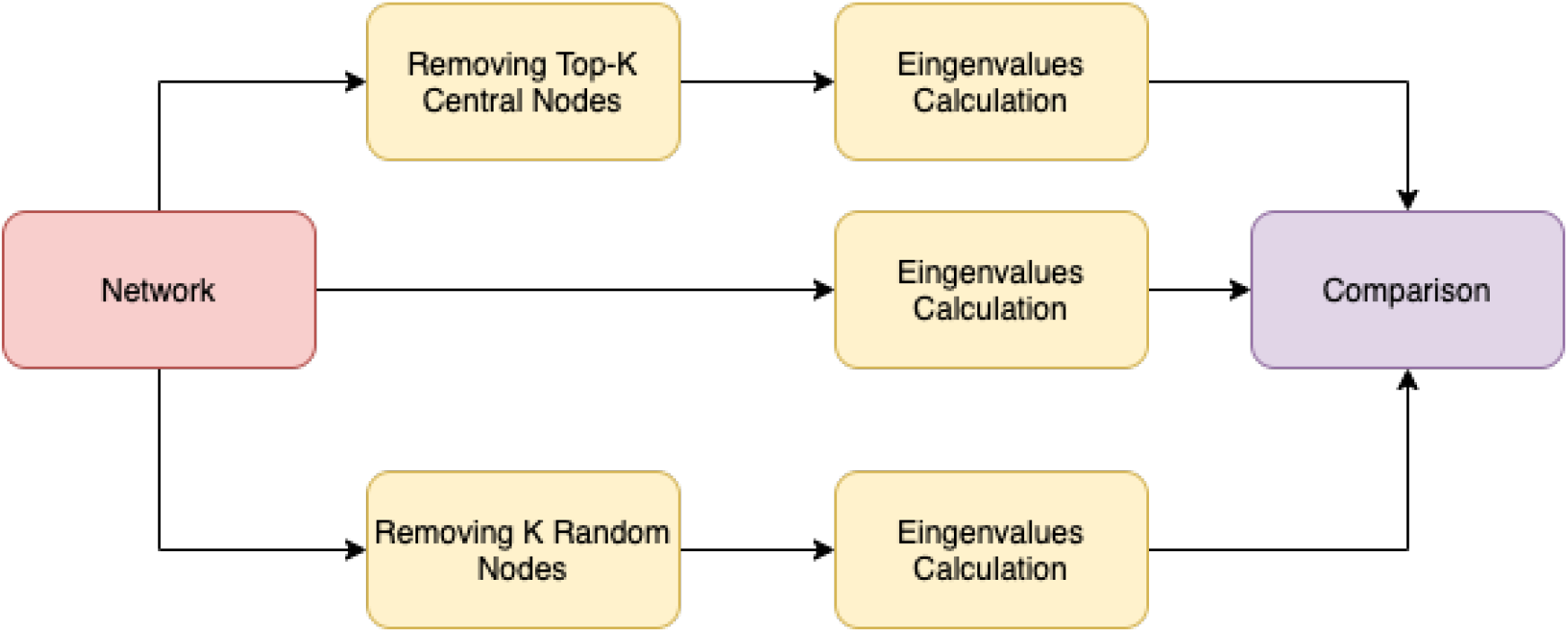
We initially built a test network. Then we determined the k most central nodes. We built two networks: one in which we removed the top-k central nodes and a second one in which we removed k randomly selected nodes. We compared the spectra of the adjacency matrices of these two networks with respect to the spectrum of the input network. We noted that the largest eigenvalues of the adjacency matrix had a more significant decrease when considering the removal of central nodes.

Consequently, we retain that any strategy of optimization that is unaware of virus spreading topology without severe lockdown may fail in virus circulation mitigation. This may favorite the insurgence of novel virus variants during the vaccination that are not covered by existing vaccines.

## 2 Materials and Methods

We used the NetworkX package for Python ver. 3.9. We initially built a set of synthetic networks using different graph random models: Erdős–Rényi, Duplication Divergence, and Barabási-Albert. Each node of the graph represented a person, while an edge modeled a contact between them. We have simulated the effect of the vaccination by eliminating the nodes representing the vaccinated individuals from the graph. The resulting graph was then an SIS model of the infection. We considered the *robustness* of a graph by calculating the spreading parameter given by the equation 3.

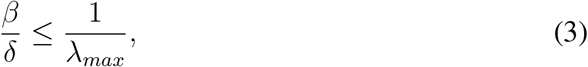

where *λ*_*max*_ is the largest eigenvalue of the adjacency matrix. Consequently, given the property that:

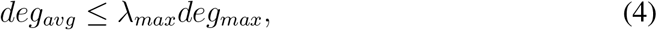

where *deg*_*avg*_ and *degsub*_*max*_ are respectively the average and the maximum node degree.

We simulated two types of vaccination strategies: topology-based, which prioritizes the most central nodes, and random (similar to the age priority strategy).

## 3 Results

Table 3 summarizes our results. The table shows for each random graph model the largest eigenvalue of the adjacency matrix of the graph considering, respectively, the original graph, the graph obtained after removing the top-k (k=100) most central nodes, and the graph obtained by removing k random nodes. We here report also the statistical significance of the difference between the decrease obtained by the removal of top-k nodes and random nodes. The analysis reported similar values for each measure: degree, betweenness, and eigenvalue centrality. In all cases, the decrease obtained when removing central nodes was significantly higher. Therefore the impact on the ratio

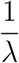

was significantly higher.

## 4 Conclusion

COVID-19 infection is currently spreading at high rates, and the current criteria for prioritizing vaccinations should be revisited to minimize the impact of the disease. We modeled the viral spreading using a network-based approach, where nodes represented subjects while the edges represented their social contacts. In such a model, it is already known that spreading may be efficiently stopped by disconnecting the network, i.e., vaccinating the most *central* or relevant nodes, therefore, eliminating “bridge edges.” We demonstrated that a topology-aware strategy may outperform, in some cases, an age-based vaccination strategy.

## Data Availability

All data produced in the present study are available upon reasonable request to the authors

---

**Table 1:** Results. The table shows the value of the largest eigenvalue (expressed as mean and standard deviation). For each random graph model and for each considered centrality value, we reported the value of the largest eigenvalue on the original graph (Original Matrix column), the value after deleting the top-k central nodes (column Top-K Matrix), the value after deleting k randomly selected nodes (Randomly Column), and the p-values of significance. DC stands for *degree centrality*, BC for *betweenness centrality*, EC for *eigenvalue centrality*, while CM stands for *Centrality Measure*. RND stands for *random*. A p-value lower than 0.05 means that the decrease of the magnitude of the largest eigenvalue was significantly higher after deleting the Top-K central nodes.

| Graph Model | CM | Original Matrix | Top-K Matrix | Randomly | p-value |
| --- | --- | --- | --- | --- | --- |
| Erdős-Rényi | DC | $400 \pm 0.51$ | $351.2 \pm 0.65$ | $368 \pm 0.7$ | $\leq 0.01$ |
| Erdős-Rényi | BC | $400 \pm 0.51$ | $354.2 \pm 0.65$ | $365 \pm 0.7$ | $\leq 0.01$ |
| Erdős-Rényi | EC | $400 \pm 0.51$ | $351.4 \pm 0.21$ | $367 \pm 0.92$ | $\leq 0.01$ |
| $G_{n,p}$ RND | DC | $400.18 \pm 0.66$ | $354.59 \pm 0.61$ | $365.44 \pm 0.62$ | $\leq 0.01$ |
| $G_{n,p}$ RND | BC | $400.18 \pm 0.66$ | $357.79 \pm 0.61$ | $368.44 \pm 0.62$ | $\leq 0.01$ |
| $G_{n,p}$ RND | EC | $400.18 \pm 0.66$ | $351.23 \pm 0.21$ | $371.44 \pm 0.62$ | $\leq 0.01$ |
| Duplication Divergence | DC | $15.2 \pm 1.81$ | $4.64 \pm 0.52$ | $15.08 \pm 1.79$ | $\leq 0.01$ |
| Duplication Divergence | BC | $15.2 \pm 1.81$ | $5.67 \pm 0.51$ | $14.88 \pm 1.19$ | $\leq 0.01$ |
| Duplication Divergence | EC | $15.2 \pm 1.81$ | $8.53 \pm 1.4$ | $17.14 \pm 1.14$ | $\leq 0.01$ |
| Barabasi-Albert | DC | $129.38 \pm 1.9$ | $67.43 \pm 3.45$ | $115.85 \pm 1.79$ | $\leq 0.01$ |
| Barabasi-Albert | BC | $129.38 \pm 1.81$ | $65.79 \pm 0.51$ | $120.98 \pm 1.19$ | $\leq 0.01$ |
| Barabasi-Albert | EC | $129.38 \pm 1.9$ | $68.53 \pm 1.4$ | $117.14 \pm 1.14$ | $\leq 0.01$ |

## References

[1] Rasim Alguliyev, Ramiz Aliguliyev, and Farhad Yusifov. Graph modelling for tracking the covid-19 pandemic spread. Infectious Disease Modelling, 6:112–122, 2021.

[2] Caroline Ash. Superspreaders are local and disproportionate. Science, 355(6329):1036–1036, 2017.

[3] Stephen P. Borgatti. Identifying sets of key players in a social network. Computational & Mathematical Organization Theory, (12):21–34, 2006.

[4] Kate M Bubar, Kyle Reinholt, Stephen M Kissler, Marc Lipsitch, Sarah Cobey, Yonatan H Grad, and Daniel B Larremore. Model-informed covid-19 vaccine prioritization strategies by age and serostatus. Science, 371(6532):916–921, 2021.

[5] Jack H Buckner, Gerardo Chowell, and Michael R Springborn. Dynamic prioritization of covid-19 vaccines when social distancing is limited for essential workers. Proceedings of the National Academy of Sciences, 118(16), 2021.

[6] Carlo Vittorio Cannistraci, Maria Grazia Valsecchi, and Ilaria Capua. Age-sex population adjusted analysis of disease severity in epidemics as a tool to devise public health policies for covid-19. Scientific reports, 11(1):1–8, 2021.

[7] Johnah C Galicia, Pietro H Guzzi, Federico M Giorgi, and Asma A Khan. Predicting the response of the dental pulp to sars-cov2 infection: a transcriptome-wide effect cross-analysis. Genes & Immunity, 21(5):360–363, 2020.

[8] Giulia Giordano, Franco Blanchini, Raffaele Bruno, Patrizio Colaneri, Alessandro Di Filippo, Angela Di Matteo, and Marta Colaneri. Modelling the covid-19 epidemic and implementation of population-wide interventions in italy. Nature medicine, 26(6):855–860, 2020.

[9] Joshua R Goldstein, Thomas Cassidy, and Kenneth W Wachter. Vaccinating the oldest against covid-19 saves both the most lives and most years of life. Proceedings of the National Academy of Sciences, 118(11), 2021.

[10] Lawrence O Gostin. The coronavirus pandemic 1 year on—what went wrongã JAMA, 325(12):1132–1133, 2021.

[11] Yan-Rong Guo, Qing-Dong Cao, Zhong-Si Hong, Yuan-Yang Tan, Shou-Deng Chen, Hong-Jun Jin, Kai-Sen Tan, De-Yun Wang, and Yan Yan. The origin, transmission and clinical therapies on coronavirus disease 2019 (covid-19) outbreak–an update on the status. Military Medical Research, 7(1):1–10, 2020.

[12] Pietro H Guzzi, Daniele Mercatelli, Carmine Ceraolo, and Federico M Giorgi. Master regulator analysis of the sars-cov-2/human interactome. Journal of clinical medicine, 9(4):982, 2020.

[13] Peter C Jentsch, Madhur Anand, and Chris T Bauch. Prioritising covid-19 vaccination in changing social and epidemiological landscapes: a mathematical modelling study. The Lancet Infectious Diseases, 2021.

[14] Jayanta Kumar Das, Giuseppe Tradigo, Pierangelo Veltri, Pietro H Guzzi, and Swarup Roy. Data science in unveiling covid-19 pathogenesis and diagnosis: evolutionary origin to drug repurposing. Briefings in Bioinformatics, 22(2):855–872, 2021.

[15] T Thanh Le, Zacharias Andreadakis, Arun Kumar, R Góez Román, Stig Tollefsen, Melanie Saville, Stephen Mayhew, et al. The covid-19 vaccine development landscape. Nat Rev Drug Discov, 19(5):305–306, 2020.

[16] Parul Maheshwari and Réka Albert. Network model and analysis of the spread of covid-19 with social distancing. Applied network science, 5(1):1–13, 2020.

[17] Francesco Ortuso, Daniele Mercatelli, Pietro Hiram Guzzi, and Federico Manuel Giorgi. Structural genetics of circulating variants affecting the sars-cov-2 spike/human ace2 complex. Journal of Biomolecular Structure and Dynamics, pages 1–11, 2021.

[18] Robert Paluch, Xiaoyan Lu, Krzysztof Suchecki, Bolesław K Szymański, and Janusz A Hołyst. Fast and accurate detection of spread source in large complex networks. Scientific reports, 8(1):1–10, 2018.

[19] Rohan Patil, Raviraj Dave, Harsh Patel, Viraj M Shah, Deep Chakrabarti, and Udit Bhatia. Assessing the interplay between travel patterns and sars-cov-2 outbreak in realistic urban setting. Applied Network Science, 6(1):1–19, 2021.

[20] Clara Stegehuis, Remco Van Der Hofstad, and Johan SH Van Leeuwaarden. Epidemic spreading on complex networks with community structures. Scientific reports, 6(1):1–7, 2016.

[21] Yang Wang, Deepayan Chakrabarti, Chenxi Wang, and Christos Faloutsos. Epidemic spreading in real networks: An eigenvalue viewpoint. In 22nd International Symposium on Reliable Distributed Systems, 2003. Proceedings., pages 25–34. IEEE, 2003.

